# Left Atrioventricular Coupling Index in Heart Failure Patients Using Echocardiography: A Simple Yet Effective Metric

**DOI:** 10.1101/2024.06.27.24309616

**Authors:** Hai Nguyen Ngoc Dang, Thang Viet Luong, Hung Khanh Tran, Ny Ha Tuyet Le, Minh Hoang Nhat Nguyen, Thang Chi Doan, Hung Minh Nguyen

## Abstract

**Background:** Heart failure (HF) is a global health challenge, with significant rates of hospitalization and mortality. Timely diagnosis is crucial for patient outcomes. Noninvasive imaging techniques assess early changes in HF but typically focus on specific aspects of the left atrium (LA) or left ventricle (LV). However, the close physiological interplay between the LA and LV suggests that indices evaluating both chambers, like the left atrioventricular coupling index (LACI), may offer superior diagnosis value for HF.

**Method:** The cross-sectional study randomly selected 1145 people at a hospital in Vietnam. Following the exclusion criteria, 160 subjects were eligible for analysis and divided into patient and healthy control groups. The patient group consisted of 60 adults meeting the criteria for HF diagnosis according to the 2022 AHA/ACC/HFSA Guideline.

**Results:** LACI levels were significantly higher in the HF group compared to the control group. Notably, LACI levels were higher in the HF with preserved ejection fraction (HFpEF) group than in the HF with reduced ejection fraction (HFrEF) group. The LACI showed a diagnostic value for HFpEF, with the highest AUC of 0.95 (95% CI: 0.921 - 0.981, p<0.001) compared to LASr, LAScd, and LAVI, with an optimal threshold of 33.07 (sensitivity: 97.1%, specificity: 87.3%). In multivariate analysis, LACI was an independent factor of HFpEF when compared to standard indices for diagnosing HFpEF (OR = 1.144, 95% CI: 1.087–1.205, p<0.001).

**Conclusion:** HF patients display increased LACI variability compared to healthy individuals, with the most significant increase observed in the HFpEF group. The LACI is an easily assessable parameter with potential value in diagnosing HFpEF.

**What is already known on this topic:** Previous studies assessing the Left Atrioventricular Coupling Index (LACI) using cardiac magnetic resonance imaging have demonstrated its predictive role in cardiovascular risk. However, clinical practice shows that echocardiography can also calculate the LACI. Echocardiography is widely available and can be performed repeatedly. Despite its promising utility, no studies to date have utilized echocardiography-measured LACI for diagnosing heart failure with preserved ejection fraction (HFpEF).

**What this study adds:** Patients with heart failure exhibit significantly higher LACI compared to the control group. Among heart failure patients, HFpEF have the highest LACI values. Additionally, LACI demonstrates superior diagnostic value for HFpEF compared to current echocardiographic parameters recommended in clinical practice.

**How this study might affect research, practice or policy:** This study holds promising potential by contributing to the development of a straightforward echocardiographic index that can diagnose HFpEF compared to current measures. Further research on LACI across diverse global populations could provide more compelling evidence of its efficacy. This could potentially lead to recommendations for integrating this index into clinical practice guidelines for guiding the clinical diagnosis of HFpEF.

**Graphical Abstract:** Graphical Abstract: Left Atrioventricular Coupling Index in Heart Failure Patients Using Echocardiography: A Simple Yet Effective Metric. HFpEF, heart failure with preserved ejection fraction; LACI, left atrioventricular coupling index; LAVI, left atrial volume index, LASr, left atrial reservoir strain, LV GLS, left ventricular global longitudinal strain; LV EF, left ventricular ejection fraction; LAEDV, left atrial end-diastolic volume; LVEDV, left ventricular end-diastolic volume.

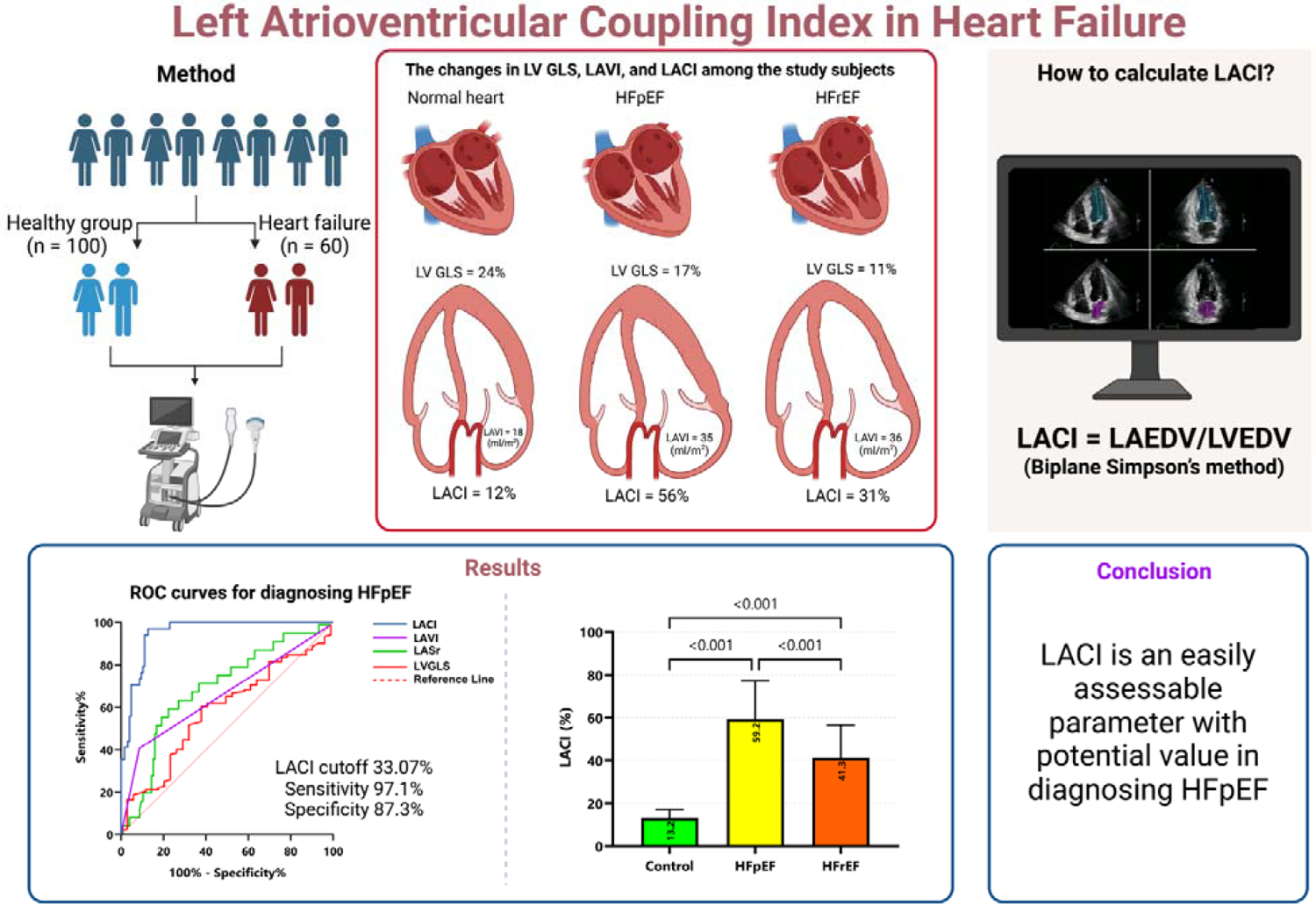

## INTRODUCTION

Heart failure (HF) is considered the end stage of various cardiovascular pathologies, such as hypertension, coronary artery disease, valvular diseases, and structural heart diseases. It poses a significant health challenge with high rates of hospitalization and mortality worldwide, including in Vietnam.^1–4^ Estimates suggest that approximately 64 million people suffer from HF globally, with a rising trend attributed partly to population aging.^2^ In patients with systolic HF, the 5-year survival rate is approximately 50%, dropping to 35% after ten years.^5^ This imposes a substantial economic burden on both patients and communities.^6^

Given this scenario, early diagnosis is deemed crucial because it can enhance both the quality of life and longevity of patients.^7^ Studies have utilized noninvasive imaging to assess structural and functional alterations of the left ventricle (LV) and left atrium (LA) early to identify individuals at high risk of HF. Parameters such as the LV ejection fraction (LV EF), LV mass index (LVMI), global longitudinal strain of the LV (LV GLS), or LA volume index (LAVI) and LA strain have shown diagnostic value in HF, particularly in heart failure preserved ejection fraction (HFpEF).^8–12^ However, these findings solely reflect individual structural/functional aspects of the LA or LV. At the same time, the pathophysiology of HF has demonstrated a close physiological relationship between the LA and LV.^13^ This raises the hypothesis that an index assessing both the LA and LV simultaneously could better predict HF than other individual indices of the LA or LV.

Recent study has further supported this. This study has shown that the left atrioventricular coupling index (LACI), calculated as the LA end-diastolic volume divided by the LV end-diastolic volume using cardiac magnetic resonance imaging, is a prognostic indicator of cardiovascular events over ten years.^14^ Although cardiac magnetic resonance imaging is the gold standard for assessing chamber volumes, the clinical reality in developing countries such as Vietnam limits patient access to cardiac magnetic resonance imaging compared to noninvasive modalities such as echocardiography. Additionally, in Vietnam, the characteristics of LACI among different HF patient groups and its prognostic and diagnostic value have not been investigated. Therefore, we conducted this study to examine the correlation between the LACI and other echocardiographic indices and to investigate the value of the LACI in diagnosing HF with HFpEF.

## METHODS

### Study population

This cross-sectional study was approved by the Institutional Review Board of Hue University (No: H2021/044), and it adhered strictly to the principles outlined in the Declaration of Helsinki (2013 version). The participants were adults over 18 who consented to the study before enrollment. One thousand one hundred forty-five randomly selected patients were recruited from a convenience sample from February 2022 to June 2023 at Hue University of Medicine and Pharmacy (the flowchart of participant recruitment in the study is shown in **Figure 1**). Among them, 160 subjects who met the study criteria and were not excluded were divided into patient and healthy control groups. The patient group consisted of 60 adults who met the selection criteria for the diagnosis of HF according to the 2022 AHA/ACC/HFSA Guideline for the management of HF.^15^ The exclusion criteria included severe comorbidities, pacemaker implantation, atrial fibrillation, severe aortic valve stenosis, severe aortic valve regurgitation, severe mitral valve stenosis, severe mitral valve regurgitation, life-threatening conditions, incomplete clinical data, inadequate echocardiographic data, poor echocardiographic image quality, and incomplete image analysis results. Within the heart failure patient group, two subgroups were formed: those with an LVEF ≥ 50% (HFpEF) and those with an LVEF < 50% (HFrEF). The control group comprised 100 healthy adults attending regular check-ups for disease screening, without a history of cardiovascular disease, matched in age with the patient group. Exclusion criteria included unclear or incomplete echocardiographic images hindering accurate analysis of cardiac chamber images.

**Figure 1:**
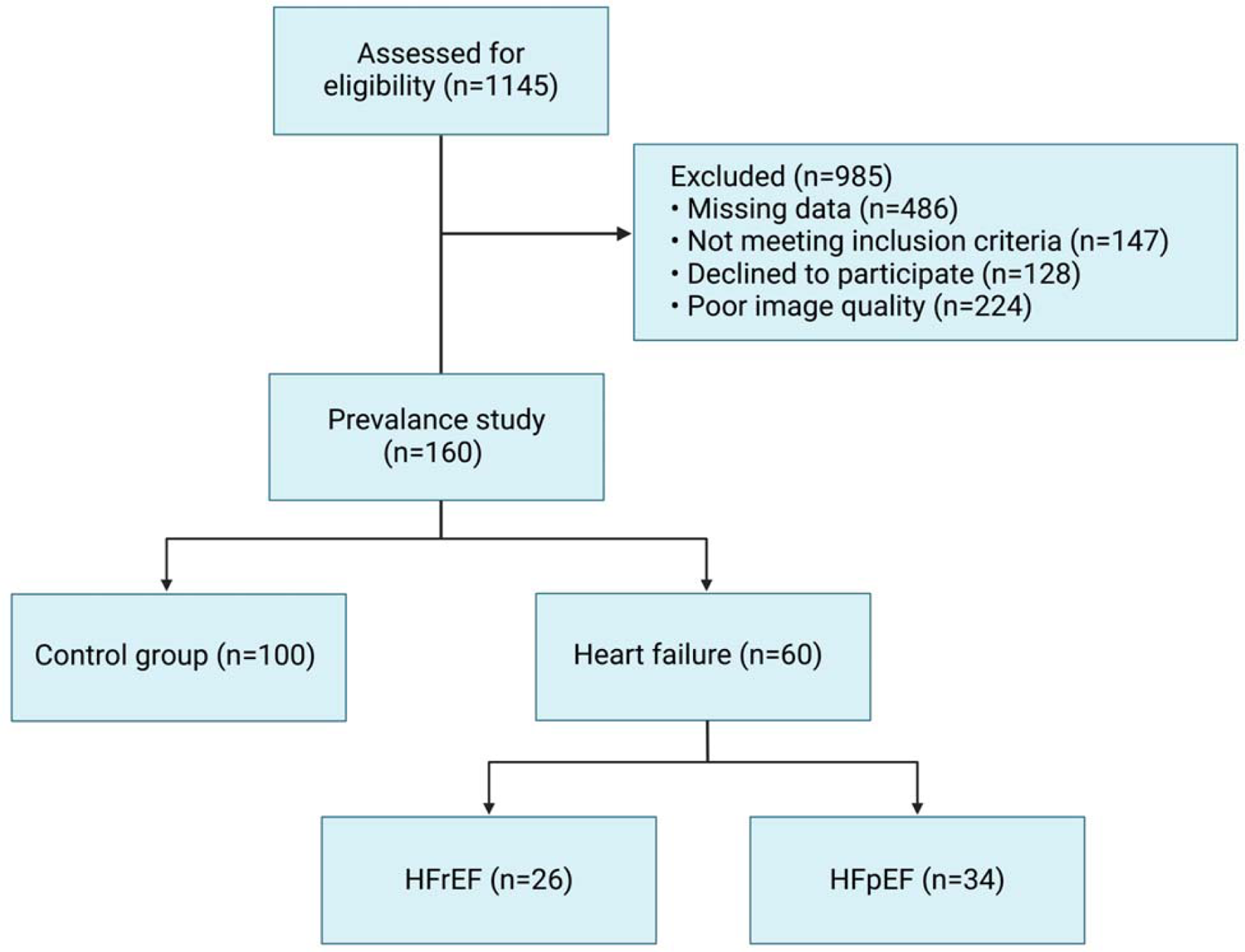
Flowchart of the selection process according to the inclusion and exclusion criteria. HFpEF, heart failure with preserved ejection fraction; HFpEF, heart failure with reduced ejection fraction.

### Clinical Data

All study participants were fully informed about the research’s benefits and risks and provided consent for data collection. Height and weight measurements were taken with a weight accuracy of 0.5 kg and a height accuracy of 1 cm. Body mass index (BMI) was calculated using the following formula: BMI = weight (kg)/[(height (m) × (height (m)]. Body surface area (BSA) was calculated using the Du Bois formula: BSA = 0.007184 × (weight)^0.425 × (height)^0.725. Blood pressure was measured according to the American Heart Association’s 2019 recommendations.^16^ Blood samples for biochemical testing were obtained from venous blood. Quantitative measurement of the N-terminal pro–B-type natriuretic peptide (NT-proBNP) concentration in the blood was performed on a Roche Cobas e 601 automated biochemical analyzer at the central laboratory unit of Hue University Hospital of Medicine and Pharmacy.

### Transthoracic echocardiography

Our study utilized a Philips Affiniti 70 ultrasound machine, which simultaneously recorded electrocardiographic signals during echocardiography. The measurement protocol followed the comprehensive transthoracic echocardiography guidelines for adults established by the American Society of Echocardiography.^17^

The myocardium was analyzed using offline QLAB software version 15 to analyze the LV GLS. All analyses were performed by a highly echocardiographer blinded to the other patient characteristics. Endocardial borders of the LV were delineated on three planes (4-chamber, 2-chamber, and 3-chamber) at end-systole when the LV size was smallest using the point-and-click technique on the LV endocardium. The software automatically performed this. In cases of incomplete tracking, manual adjustments to the endocardial borders were attempted, and if not satisfactory, the patient was excluded from the analysis. The software automatically tracked the endocardial surface of the LV, creating a region of interest adjusted to encompass the entire myocardial thickness throughout the cardiac cycle. LV GLS was calculated as the change in length divided by the original length of the speckle pattern in the cardiac cycle and expressed as a percentage, with myocardial lengthening represented as positive strain and shortening as negative strain.^18, 19^ LV GLS was calculated as the average of 18 peak systolic strain segmental results.^20^ LA strain assessment was conducted in both the two-chamber and four-chamber views, with reference points set at the onset of the P wave in the cardiac cycle. Measurements of LA strain were acquired during the reservoir, conduit, and contractile phases of LA function, designated left atrium strain reservoir function (LASr), left atrium strain conduit function (LAScd), and left atrium strain contractile function (LASct), respectively. The results of the LV and LA strains are conventionally represented as negative values. However, for convenience in analysis and display, we utilized the absolute values of these results.

### Two-Dimensional echocardiographic methods for assessment of left atrioventricular coupling index

The LACI is the ratio between the left atrial end-diastolic volume (LAEDV) and the left ventricular end-diastolic volume (LVEDV). LA and LV volumes are measured during the end-diastolic phase of the cardiac cycle, identified by the peak of the QRS complex on the electrocardiogram or by the fully closed mitral valve on echocardiography. Measurements were sequentially conducted on four-chamber and two-chamber views. Biplane Simpson’s method was employed to evaluate the LVEDV and LAEDV. The LACI value is expressed as a percentage, with higher LACI values indicating a more significant imbalance between LA and LV volumes during end-diastole, reflecting changes in left atrioventricular coupling^19, 21^. To avoid influencing the research results, the echocardiography procedure for acquiring the LACI was conducted by a separate echocardiography specialist independent of the research team. **Supplementary Figure 1** illustrates the procedure for calculating the LACI.

### Statistical analysis

All the statistical analyses were performed using IBM SPSS statistical software version 26.0, GraphPad Prism software version 10.1.2.324, and The R Project for Statistical Computing version 4.0.3. The normality of the distribution of variables was assessed using the Kolmogorov–Smirnov test. Categorical variables are presented as frequencies and percentages. Normally distributed continuous variables are expressed as the mean ± standard deviation (X ± SD) and median (interquartile range 25 and 75) if the distribution is nonnormal. The study results are presented in tables and graphs. The χ2 and Fisher’s exact tests were used to compare observed proportions and examine relationships between categorical variables. Independent samples t tests were used to compare the mean values of normally distributed continuous variables between two groups with equivalent variances. One-way ANOVA was used for comparisons involving three or more groups. The Mann‒Whitney U test was used for comparisons between two groups, and the Kruskal‒Wallis test was used for comparisons involving three or more groups when the distribution was nonnormal or variances were not equivalent. The Pearson correlation coefficient (r) between LAVI and other echocardiographic indices was assessed using Pearson correlation to determine whether the distribution was normal or Spearman correlation (r_s_) to determine whether the distribution was nonnormal. Receiver operating characteristic (ROC) curves were generated to evaluate the diagnostic value of the LACI and other echocardiographic parameters for predicting heart failure with a preserved ejection fraction (LVEF), and the area under the curve (AUC), sensitivity, and specificity were assessed. The AUC indicates the accuracy of the predictive factor, ranging from 0.5 to 1. AUC comparison was conducted using the DeLong method to assess the diagnostic value of strain compared to existing guideline criteria^22^. All the statistical tests were two-sided, and a p value < 0.05 was considered to indicate statistical significance. Logistic multivariable regression was conducted to assess the risk of HFpEF of the LACI and other echocardiographic values. We randomly selected 30 of the 160 study participants to evaluate the intraclass correlation coefficient (ICC). Intraobserver and interobserver variability of LACI was assessed using the ICC. Additionally, the Bland-Altman method assessed inter- and intraobserver reliability among echocardiography experts’ results. For intraobserver variability, the same operator independently remeasured the data after seven days. For interobserver variability, the data were reanalyzed by a second operator blinded to the initial measurements. A p value <0.05 was considered to indicate statistical significance.

## RESULTS

### Baseline demographic and clinical features of the study population

In our study, when comparing the HF group and the control group, there were no differences in age, gender, BMI, or BSA (p > 0.05). However, there were statistically significant differences in age, heart rate, systolic blood pressure, and diastolic blood pressure (p < 0.001). The detailed parameters are presented in **Table 1**.

**Table 1:**
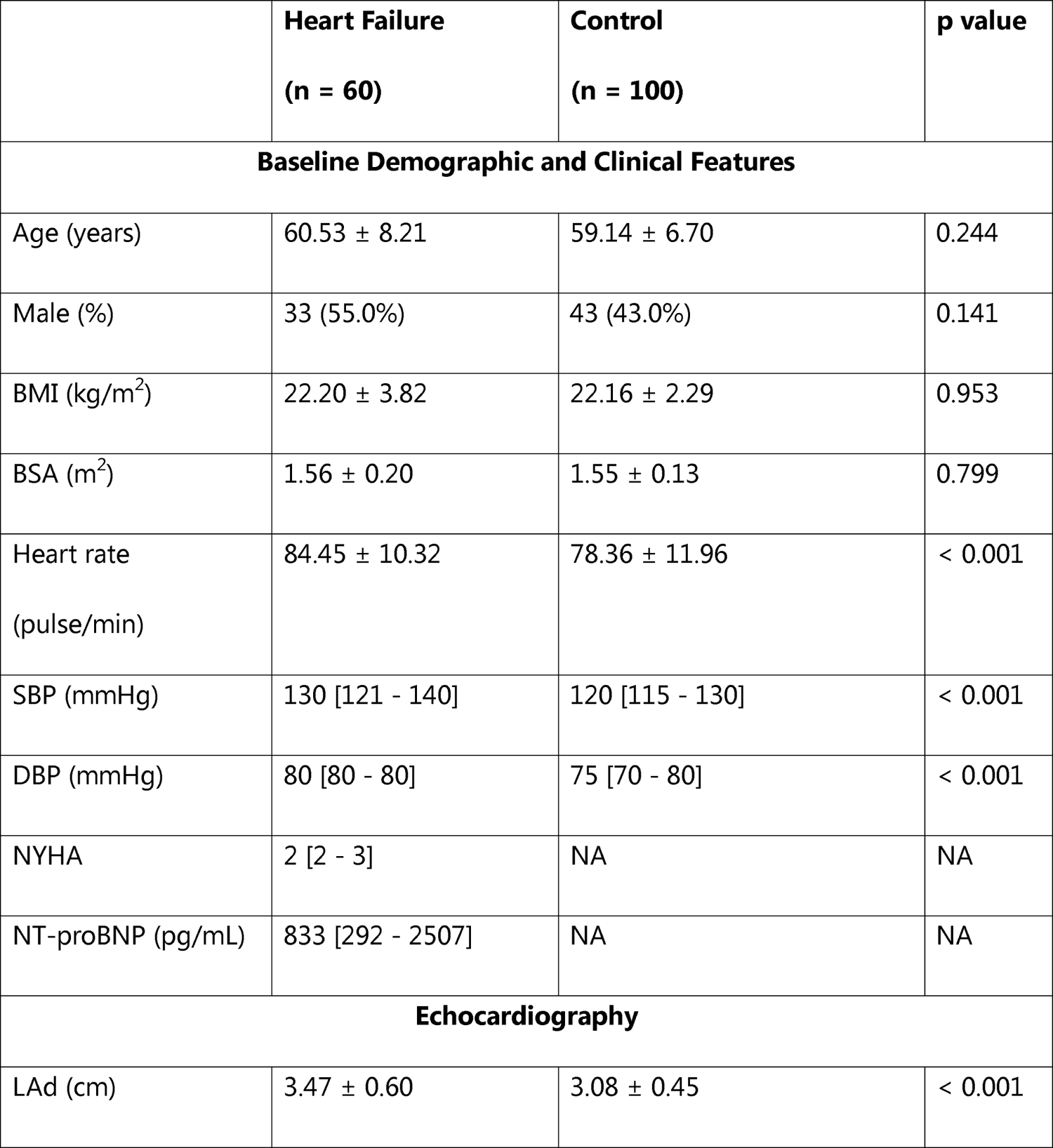
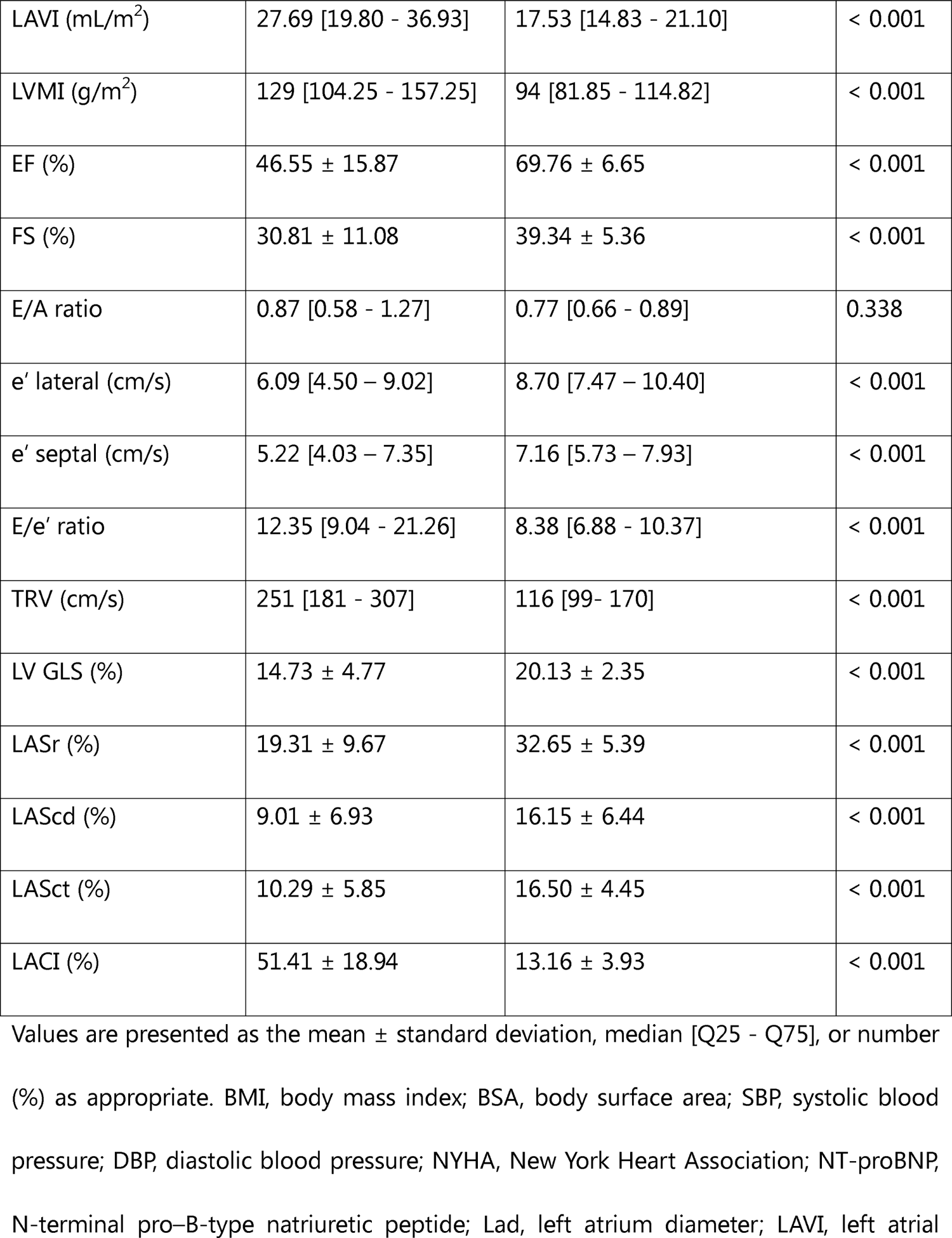
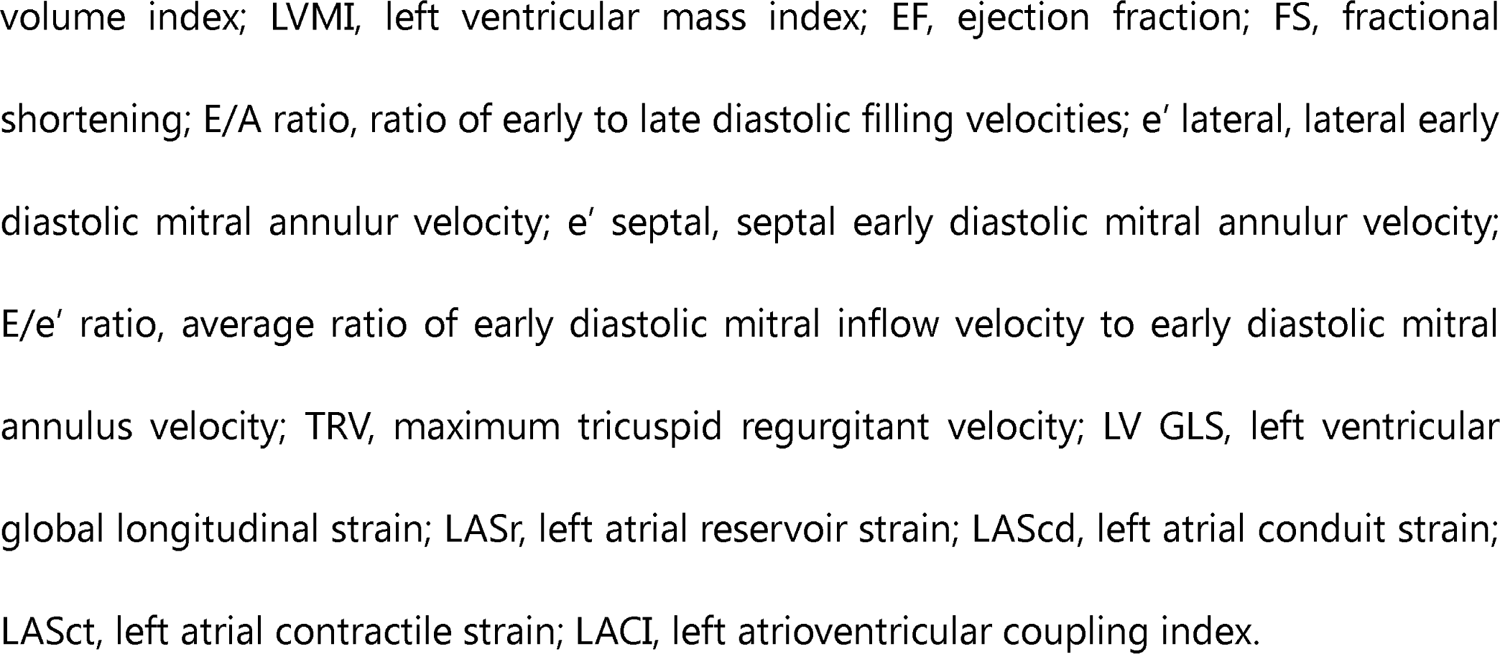
Baseline Demographic and Clinical Features and Echocardiographic Parameters of the Study Population.

In the present study, echocardiographic indices significantly differed between the control and HF groups (p < 0.001). Specifically, the LVMI and LAVI were more significant in the HF group than in the control group. Conversely, the LVEF, LV GLS, LASr, LAScd, and LASct were lower in the HF group than in the control group. Details of the echocardiographic parameters are shown in **Table 1**.

### Comparison of LACI and speckle tracking echocardiography parameters of the LA and LV in the HF and control groups

Our study revealed that the LACI in both the HFrEF and HFpEF groups was greater than that in the control group, with values of 41.28 ± 15.27% and 59.16 ± 17.94%, respectively, compared to 13.15 ± 3.92%. The LACI in the HFpEF group was significantly greater than that in the HFrEF group. The LV GLS in the HFrEF and HFpEF groups were lower than those in the control group (11.15 ± 3.29%; 17.46 ± 3.83% compared to 20.13 ± 2.34%, respectively; all p<0.001). Further details are presented in **Table 2** and illustrated in **Supplemetary Figure 2**.

**Figure 2:**
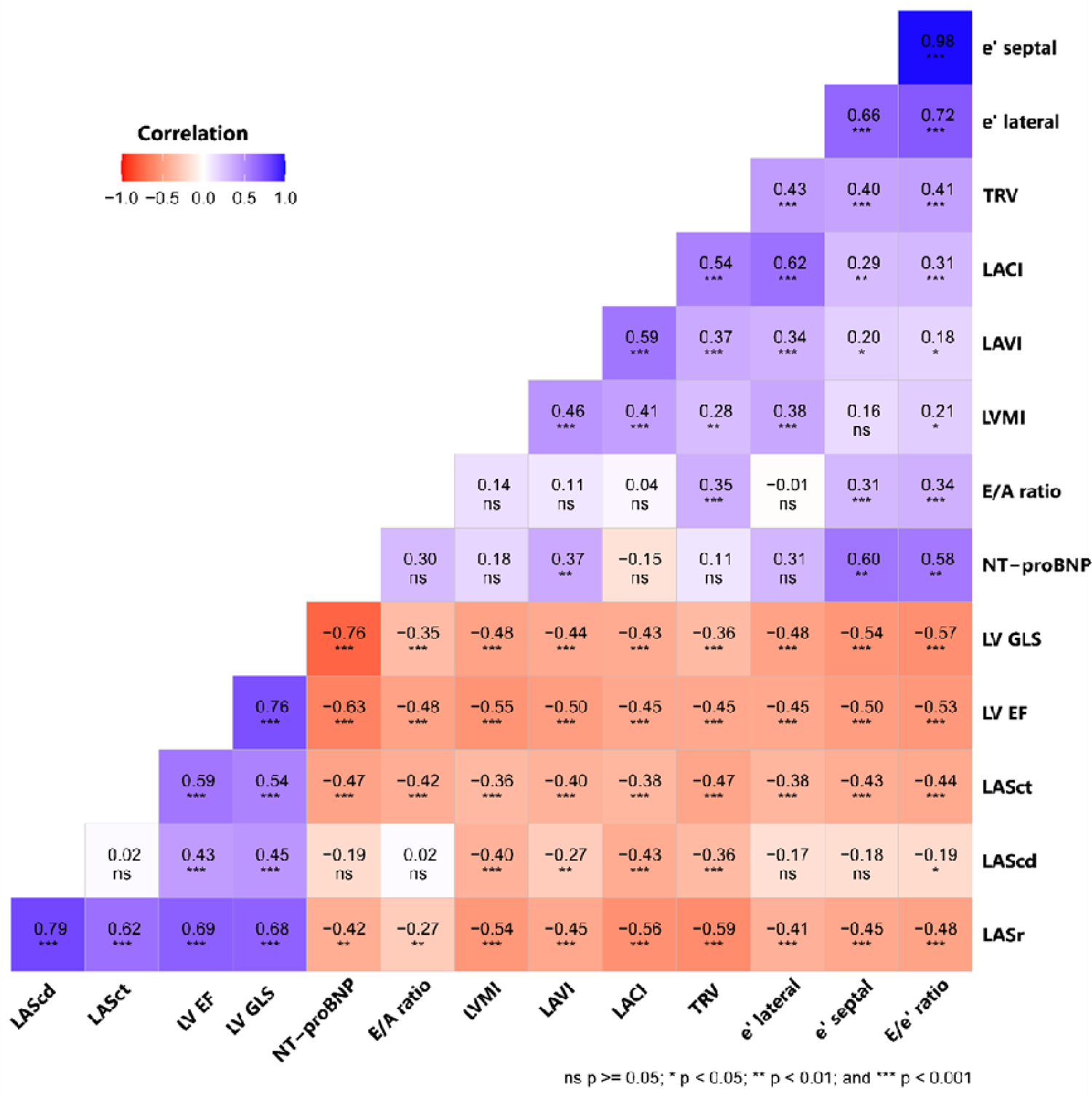
Heatmap illustrating the relationships between echocardiographic parameters. The color of each cell corresponds to the magnitude and direction of the correlation coefficient, with red indicating a positive correlation and blue indicating a negative correlation. The numerical values within the cells represent the two-way correlation coefficient. NT-proBNP, N-terminal pro–B-type natriuretic peptide; LAVI, left atrial volume index; LVMI, left ventricular mass index; LV EF, left ventricular ejection fraction; E/A, ratio of early to late diastolic filling velocities; e’ lateral, lateral early diastolic mitral annulur velocity; e’ septal, septal early diastolic mitral annulur velocity; E/e’ ratio, average ratio of early diastolic mitral inflow velocity to early diastolic mitral annulus velocity; TRV, maximum tricuspid regurgitant velocity; LV GLS, left ventricular global longitudinal strain; LASr, left atrial reservoir strain; LAScd, left atrial conduit strain; LASct, left atrial contractile strain; LACI, left atrioventricular coupling index.

**Table 2:**
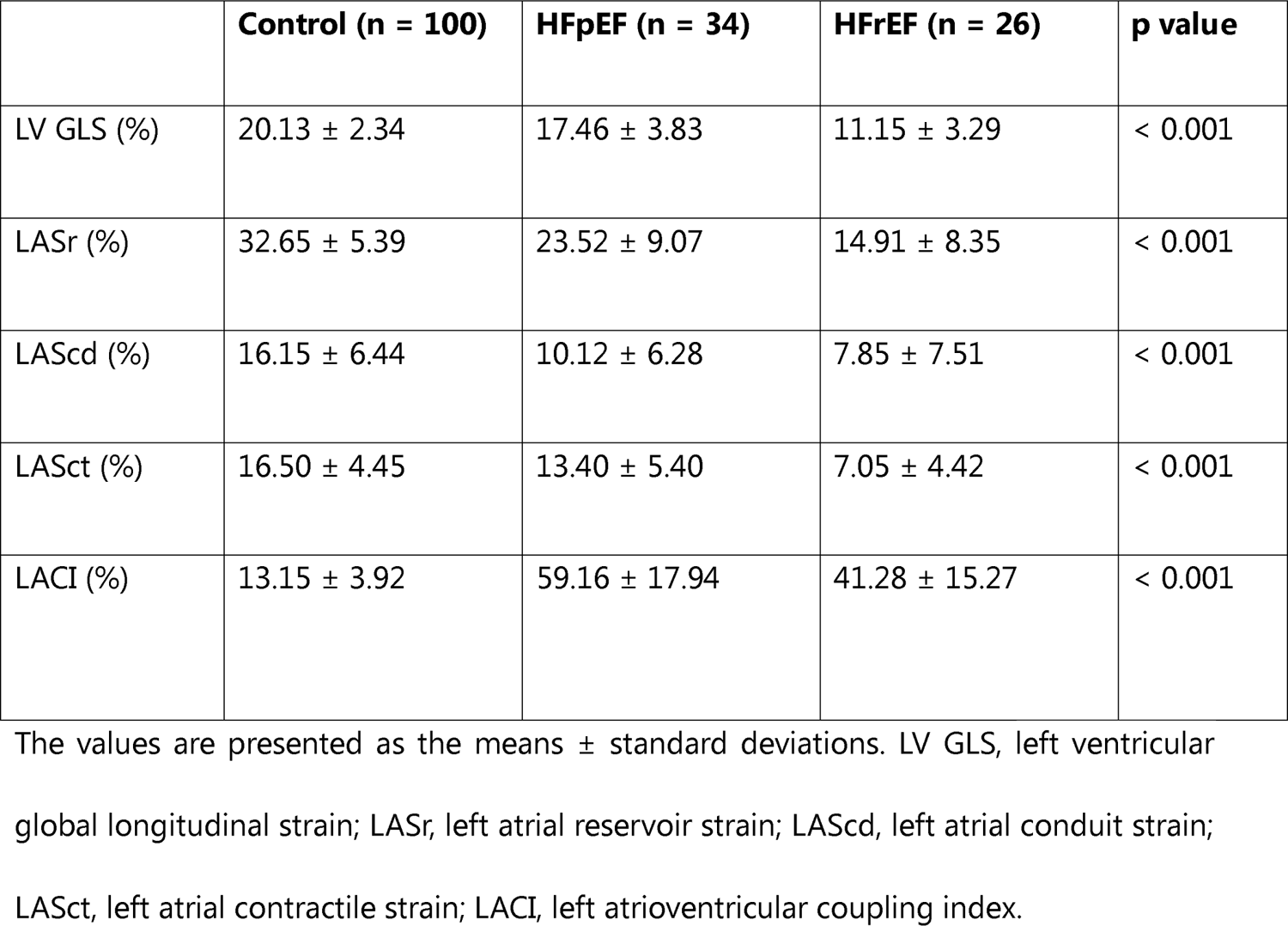
Speckle tracking echocardiography parameters of the LA, LV, and LACI of the study population.

### Correlation between LACI and other echocardiographic indices

The correlation analysis of LA and LV parameters obtained from echocardiography is visualized in **Figure 2** as a heatmap. Significant correlations were observed between LACI and EF, LAVI, LVMI, and LV GLS (EF: r = −0.45, LAVI: r_s_ = 0.59, LVMI: r_s_ = 0.41, LV GLS: r = −0.43, all p < 0.05). Further correlational details are clearly illustrated in the heatmap (**Figure 2**).

### The value of the LACI and other echocardiographic parameters in diagnosing HFpEF

The sensitivity and specificity of the LACI in diagnosing HFpEF were analyzed by constructing ROC curves. The largest area under the ROC curve (AUC) for LACI was 0.95, with an optimal threshold value of 33.07 (sensitivity 97.1%, specificity 87.3%). A statistically significant difference was observed when comparing the AUC of LACI with that of other echocardiographic parameters using the DeLong method (p < 0.05). Additionally, the other parameters’ AUC, cutoff points, sensitivity, and specificity are presented in **Table 3** and **Figure 3**.

**Figure 3:**
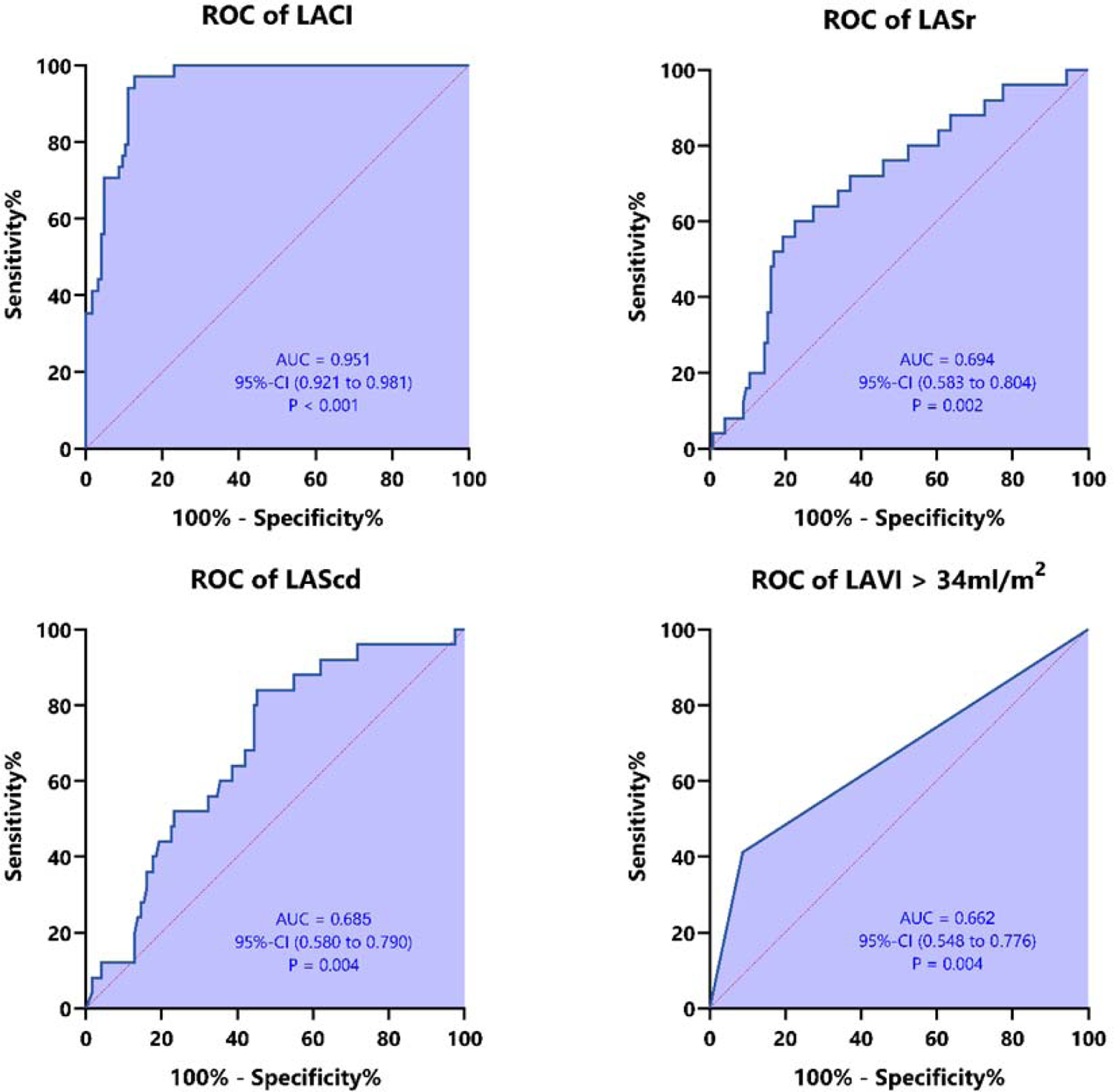
ROC curves of LASr, LAScd, LACI, and LAVI for diagnosing HFpEF. LAVI, left atrial volume index; LASr, left atrial reservoir strain; LAScd, left atrial conduit strain; LACI, left atrioventricular coupling index.

**Table 3.**
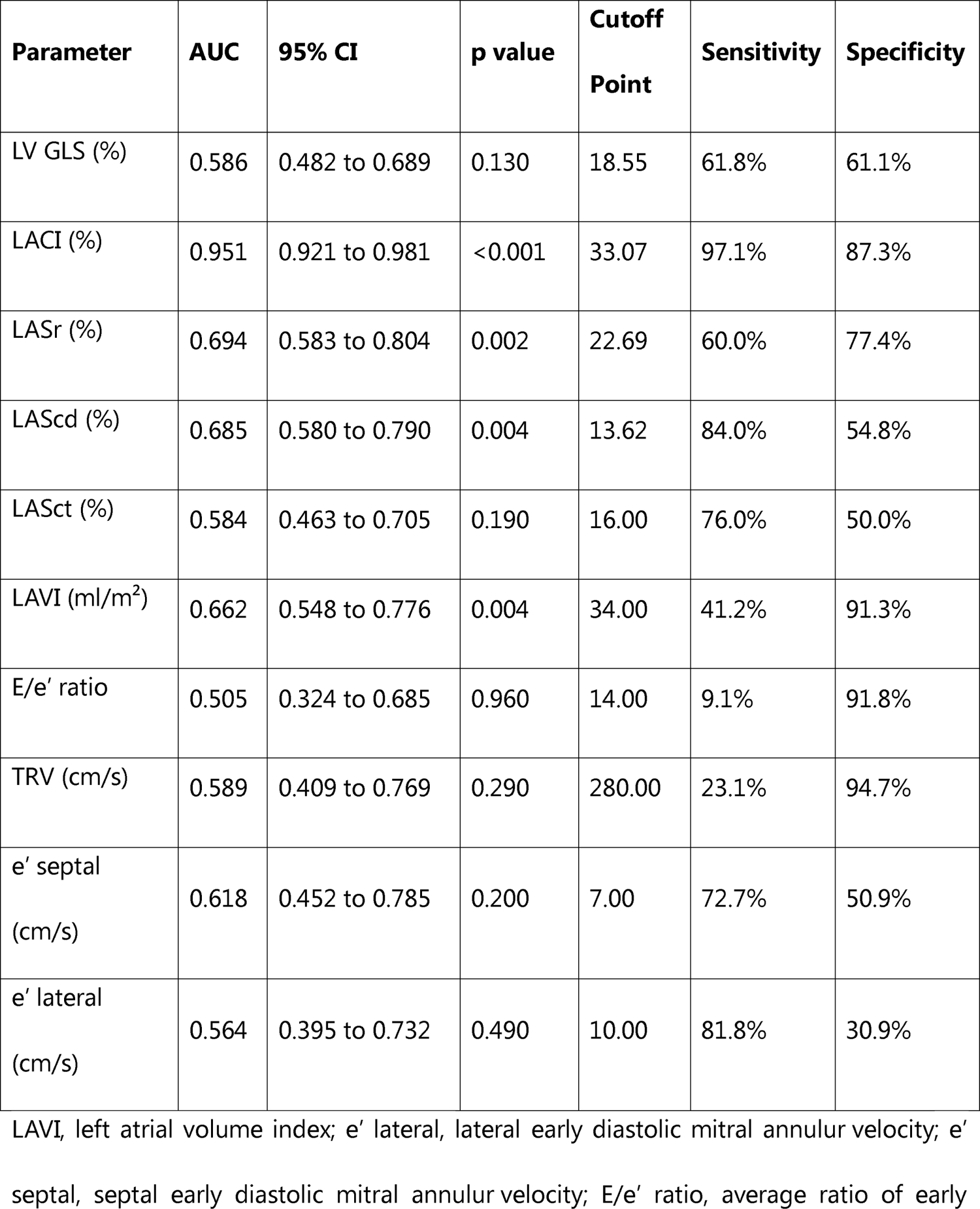
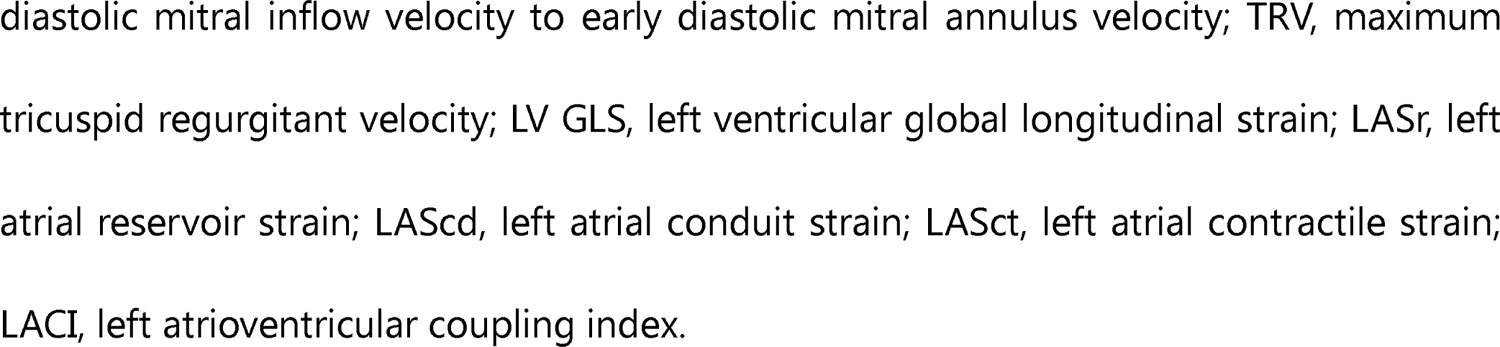
Values of echocardiographic indices in the diagnosis of HFpEF.

### Logistic regression analysis for assessing HFpEF risk factors

Logistic regression analysis was conducted to evaluate the risk factors for HFpEF using LACI and other echocardiographic indices as covariates. According to the univariate analysis, higher LACI and lower LASr were identified as risk factors for HFpEF. Furthermore, our multivariate analysis found that LACI and LASr are independent factors of HFpEF compared to standard indices for diagnosing HFpEF. Further details are provided in **Table 4**.

**Table 4.**
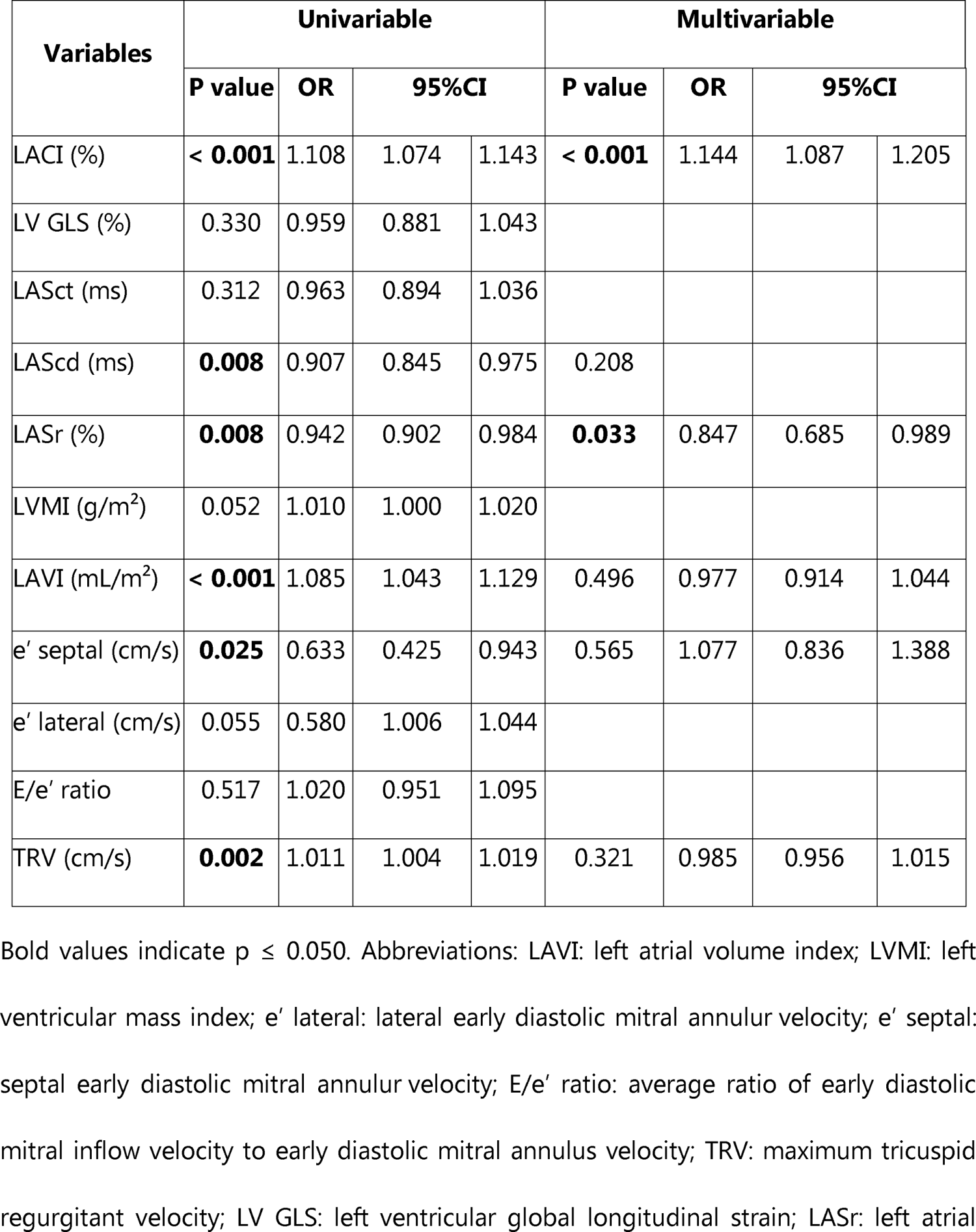
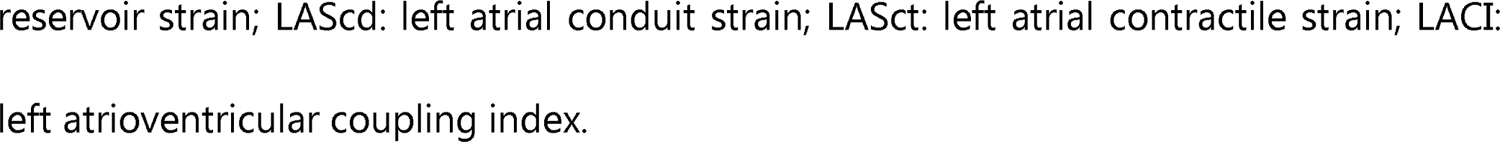
Regression analysis for prediction of HFpEF susceptibility.

### Reliability of LACI measurement

Figure 4 presents the intraobserver and interobserver variability for LACI measurements. The LACI demonstrated good reproducibility, indicated by high ICC values.

**Figure 4:**
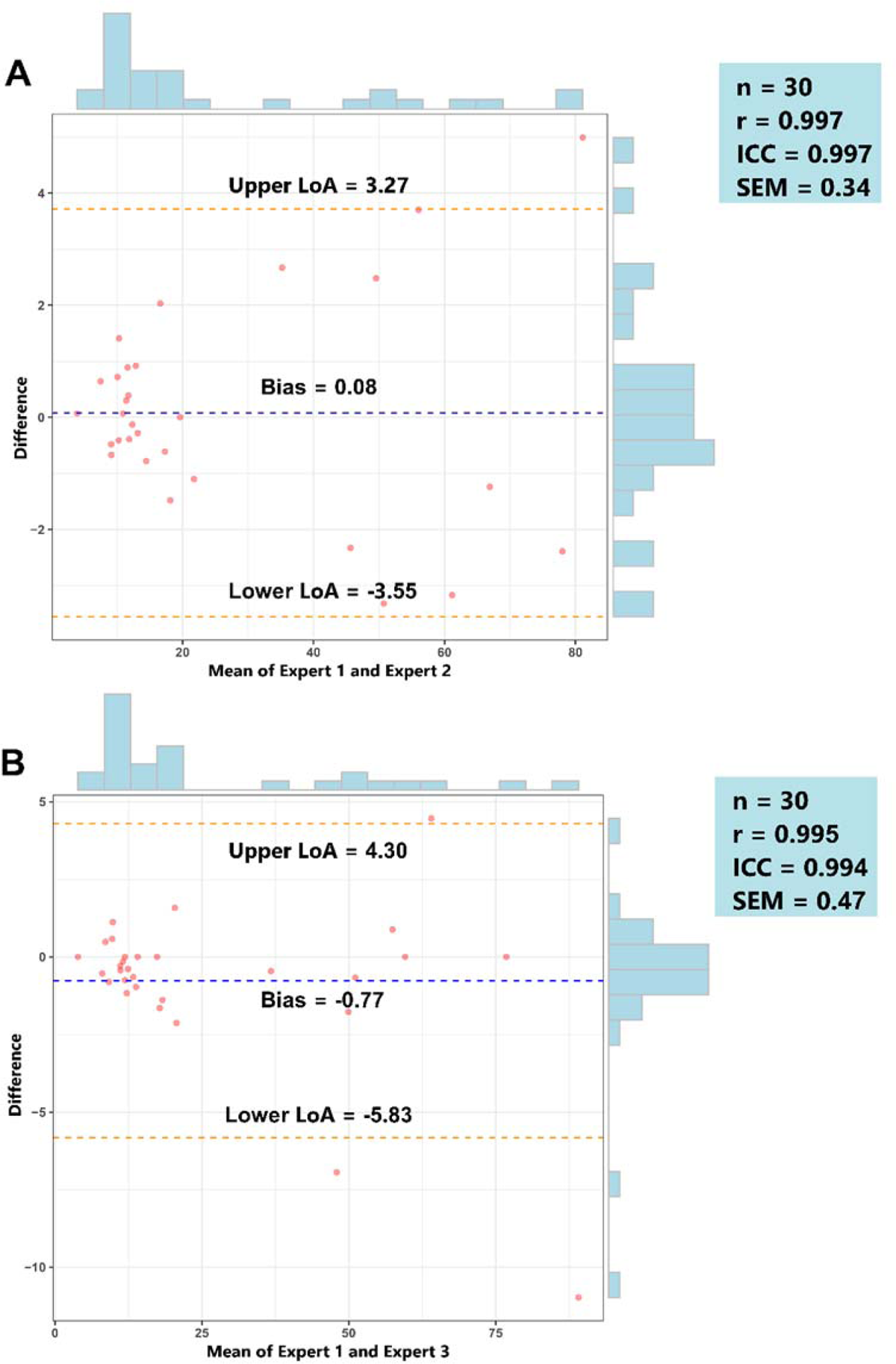
Bland-Altman plot with marginal histograms for interobserver and intraobserver agreement in assessing the reliability of LACI measurement. Panel A: Intraobserver variability of LACI. Panel B: Interobserver variability of LACI. Bias: mean of differences between the two experts, Upper LoA: upper level of limits of agreement = mean of differences + 1.96 standard deviations, Lower LoA: lower level of limits of agreement = mean of differences - 1.96 standard deviations, r: Pearson correlation coefficients, ICC: Intraclass correlation coefficient, SEM: standard error of measurement.

## DISCUSSION

Our study aimed to examine the LACI among subgroups of HF patients using echocardiography. Our findings were highly promising, as we observed significant differences in LACI among the HFpEF, HFrEF, and control groups (see **Table 2** and Figure 3). Furthermore, the LACI showed a high diagnostic value and accuracy in diagnosing HFpEF compared with commonly used indices (**Table 3** and Figure 3). Our study indicated an increase in LACI, particularly in the HFpEF group, followed by a significant decrease in the HFrEF group. The pathophysiology of HF can explain this phenomenon. In the HFpEF group, there was a characteristic increase in myocardial stiffness due to fibrosis or alterations in the extracellular matrix of the myocardial tissue, leading to impaired ventricular relaxation, particularly in the LV, resulting in diastolic dysfunction.^23, 24^ Inadequate ventricular relaxation impedes blood flow from the LA to the LV, causing increased pressure in the LA and pulmonary veins to compensate during the early diastolic phase.^25^ However, prolonged high pressure exerts more pressure on the LA wall, causing dilation of the LA, thereby increasing both the minimal and maximal LA volumes.^13^ In their study, Sung-Hee Shin and colleagues demonstrated that LAVImin is a superior index for reflecting left ventricular filling pressure and predicting HF compared to LAVImax.^26^ This is further supported by the fact that the LA bears direct pressure from the LV during end-diastole, making the LA size an optimal indicator of LV diastolic function.^27^ However, unlike changes in the LA, in HFpEF patients, the predominant change in the LV is concentric hypertrophy, leading to increased LV wall thickness but minimal impact on the LV chamber volume.^24, 28^ Epidemiological and clinical imaging studies have consistently shown that in HFpEF patients, the average LV size falls within normal limits.^28^ The disproportionate increase in LA volume relative to the LV during end-diastole results in a high LACI in HFpEF patients. Conversely, although the LACI increased in the HFrEF compared to the control group, it tended to be smaller than that in the HFpEF group. These findings are consistent because both HFrEF and HFpEF give rise to LA dilation. Specifically, eccentric LA remodeling is more pronounced in HFrEF patients, resulting in a larger LA volume than in HFpEF patients.^29^ The key difference here is the LV changes in HFrEF, characterized by diastolic dysfunction and LV dilation, eventually leading to increased LV end-diastolic volume.^24^ Consequently, the disproportionate volume between the LA and LV during end-diastole is reduced compared to that in HFpEF patients. LV diastolic and filling parameters are often used to assess cardiac function in HF patients. However, these indicators alone may not accurately reflect overall cardiac function, particularly in HFpEF patients, where LV filling parameters may not accurately reflect the severity of HF. Recent studies have highlighted the LACI as an index that can assess LV diastolic function effectively and predict HFpEF.^27, 30^ The current research demonstrated significant differences in LACI between HF patients and controls, especially in HFpEF patients. Therefore, LACI, as additional data, may be helpful for clinicians in diagnosing HFpEF.

Our study found that the LACI yielded promising results in diagnosing HFpEF. Compared to other modern techniques, such as speckle tracking echocardiography for assessing LV GLS and LA strain, which require sophisticated ultrasound machines and analysis software, widespread investment and application may be challenging, particularly in developing countries such as Vietnam. On the other hand, LACI is a simple technique that can be directly performed on 2D echocardiographic images available on most current ultrasound devices. With its favorable results and ease of implementation, LACI could be widely applied as a cost-effective technique for diagnosing HFpEF, especially in developing countries lacking resources for other advanced equipment.

## LIMITATIONS OF THE STUDY

Our study has provided valuable insights into the LACI among HF patients, especially those with HFpEF. However, certain limitations need to be acknowledged. Firstly, its small sample size and single-center focus limited the study, which may introduce selection bias. Secondly, our study on LACI did not assess patients with specific rhythm disorders, particularly atrial fibrillation, which can contribute to HF. Thirdly, this cross-sectional study can only establish correlations rather than causal relationships. Therefore, a longitudinal study is needed to address this issue. Lastly, there is a limitation regarding the research equipment, as the study was conducted using a specific ultrasound machine and ultrasound software. Further research could compare results from various manufacturers of ultrasound software.

## CONCLUSION

HF patients exhibit increased variability in LACI compared to healthy individuals, with the most significant increase observed in the HFpEF group. The LACI is an easily assessable parameter with potential value in diagnosing HFpEF.

## Data Availability

There is a restriction on sharing the dataset due to regulations from the Research Ethics Committee that approved the study and data protection laws, precisely document 13/2023/N?-CP in Vietnam. The informed consent obtained from our patients did not include a provision for third-party access to their medical information, which contains sensitive data such as date of birth, initials, date of admission, and discharge. As per the data use agreement with the Research Ethics Committee, the authors are not authorized to share or publicly release the data. The authors did not have any special access privileges to this data. Please contact Dr. Hai Nguyen Ngoc Dang, who has access to the data in some necessary cases and with appropriate licensing.

## ACKNOWLEDGMENTS

Not applicable.

## AUTHOR CONTRIBUTIONS

Hai Nguyen Ngoc Dang: Conceptualization, methodology, investigation, data curation, writing - original draft, writing - review & editing. Thang Viet Luong: Conceptualization, methodology, investigation, data curation, writing - original draft, writing - review & editing. Hung Khanh Tran, Ny Ha Tuyet Le, Minh Hoang Nhat Nguyen, Thang Chi Doan, Hung Minh Nguyen: Writing - original draft, writing - review & editing. All authors have read and agreed to the published version of the manuscript.

## DATA SHARING STATEMENT

Data cannot be shared publicly due to certain local ethical constraints. Researchers who meet the criteria for access to confidential data may contact the author, Hai Nguyen Ngoc Dang (via email at).

---

**Supplementary Figure 1:**
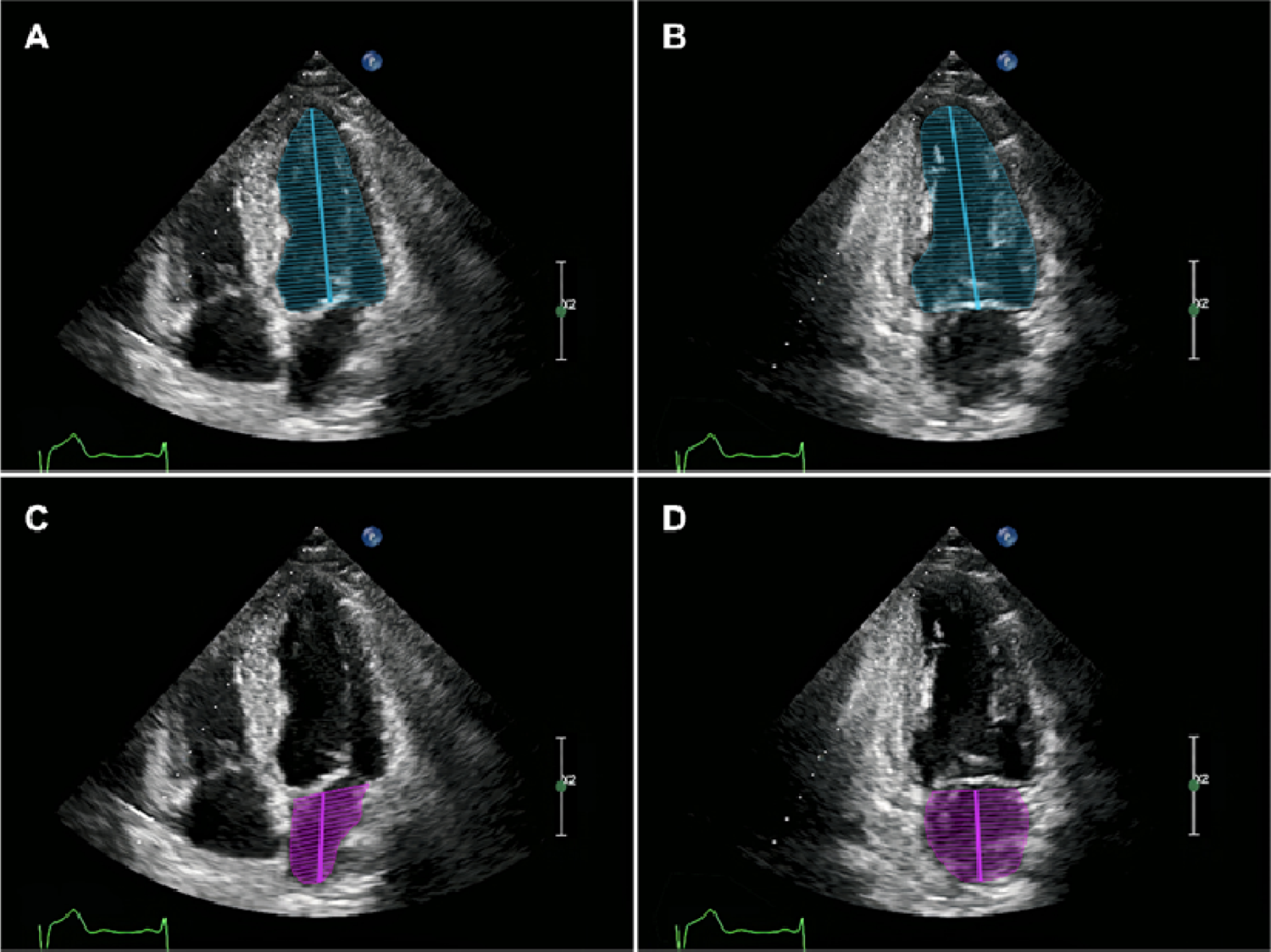
Measurement of the LAEDV and LVEDV using the biplane method to calculate the LACI index using two-dimensional echocardiography. Panel A: Measurement of LV volume in the 4-chamber view. Panel B: Measurement of LV volume in the 2-chamber view. Panel C: Measurement of LA volume in the 4-chamber view. Panel D: Measurement of LA volume in the 2-chamber view.

**Supplementary Figure 2:**
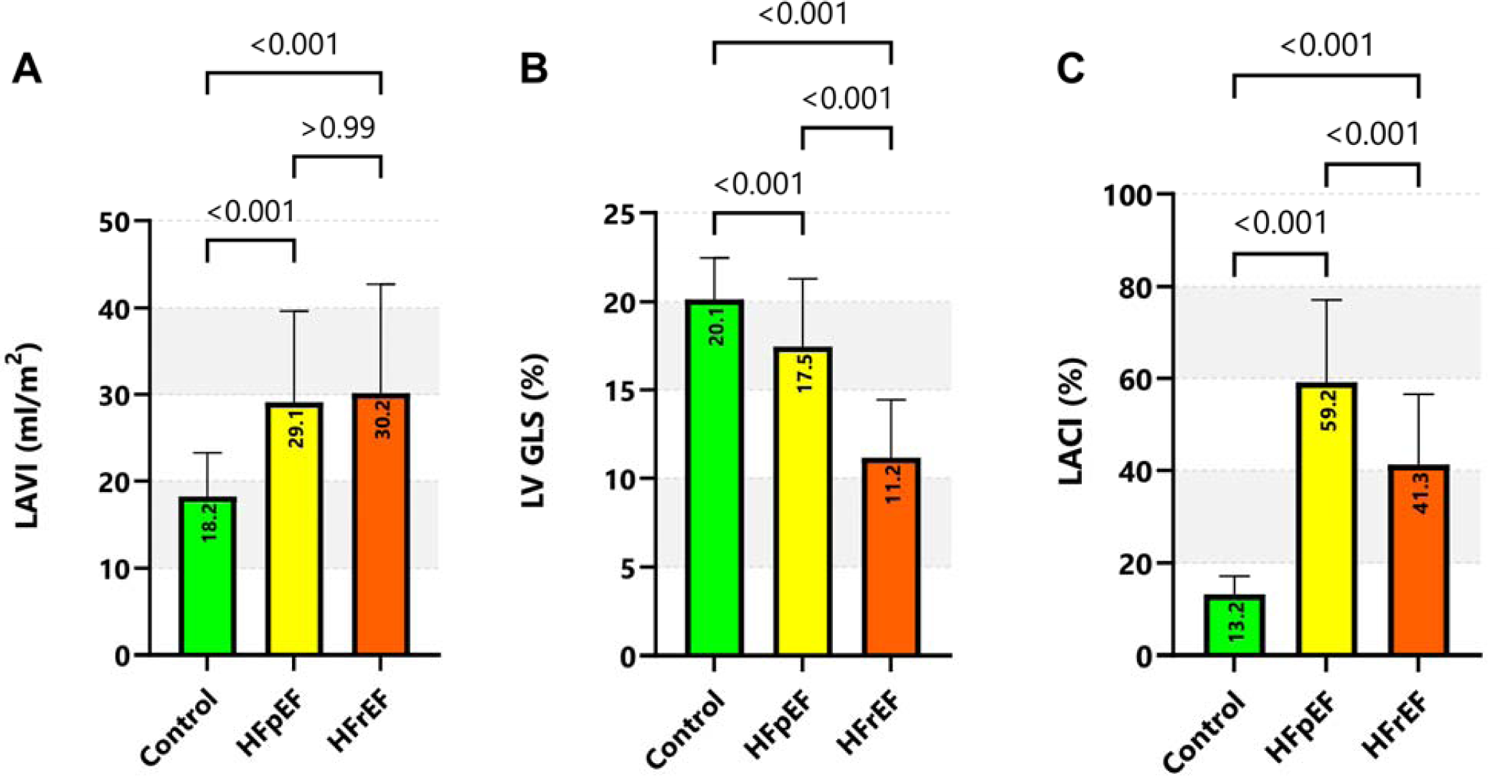
Comparison of LACI, LAVI, and LV GLS indices among HFpEF, HFrEF, and the control group. LAVI, left atrial volume index; LV GLS, global longitudinal strain; LACI, left atrioventricular coupling index; HFpEF, heart failure with preserved ejection fraction; HFpEF, heart failure with reduced ejection fraction.

