## Supplementary figures and images for "Left Atrioventricular Coupling Index in Heart Failure Patients Using Echocardiography: A Simple Yet Effective Metric"

### Supplementary Figure 1

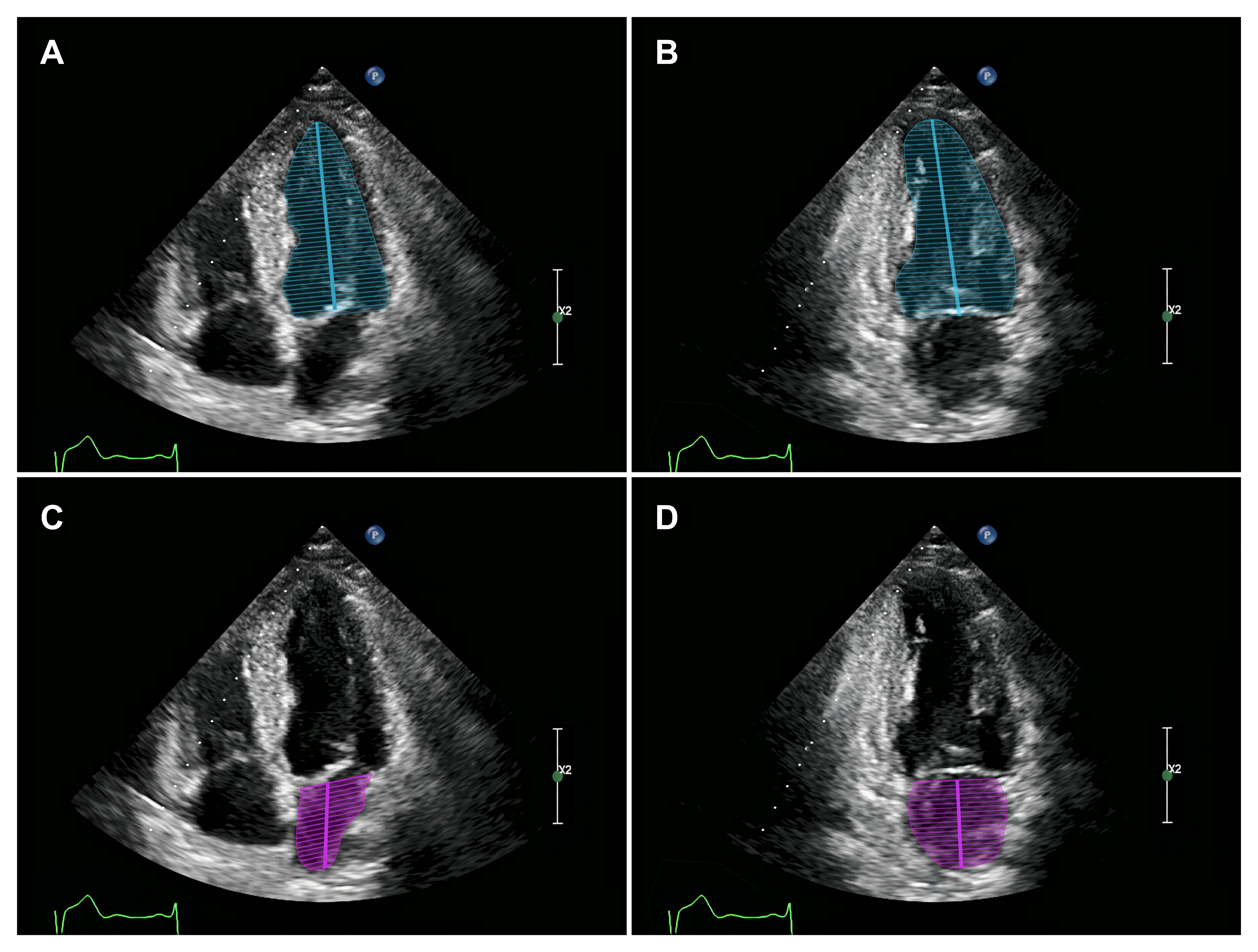

### Supplementary Figure 2

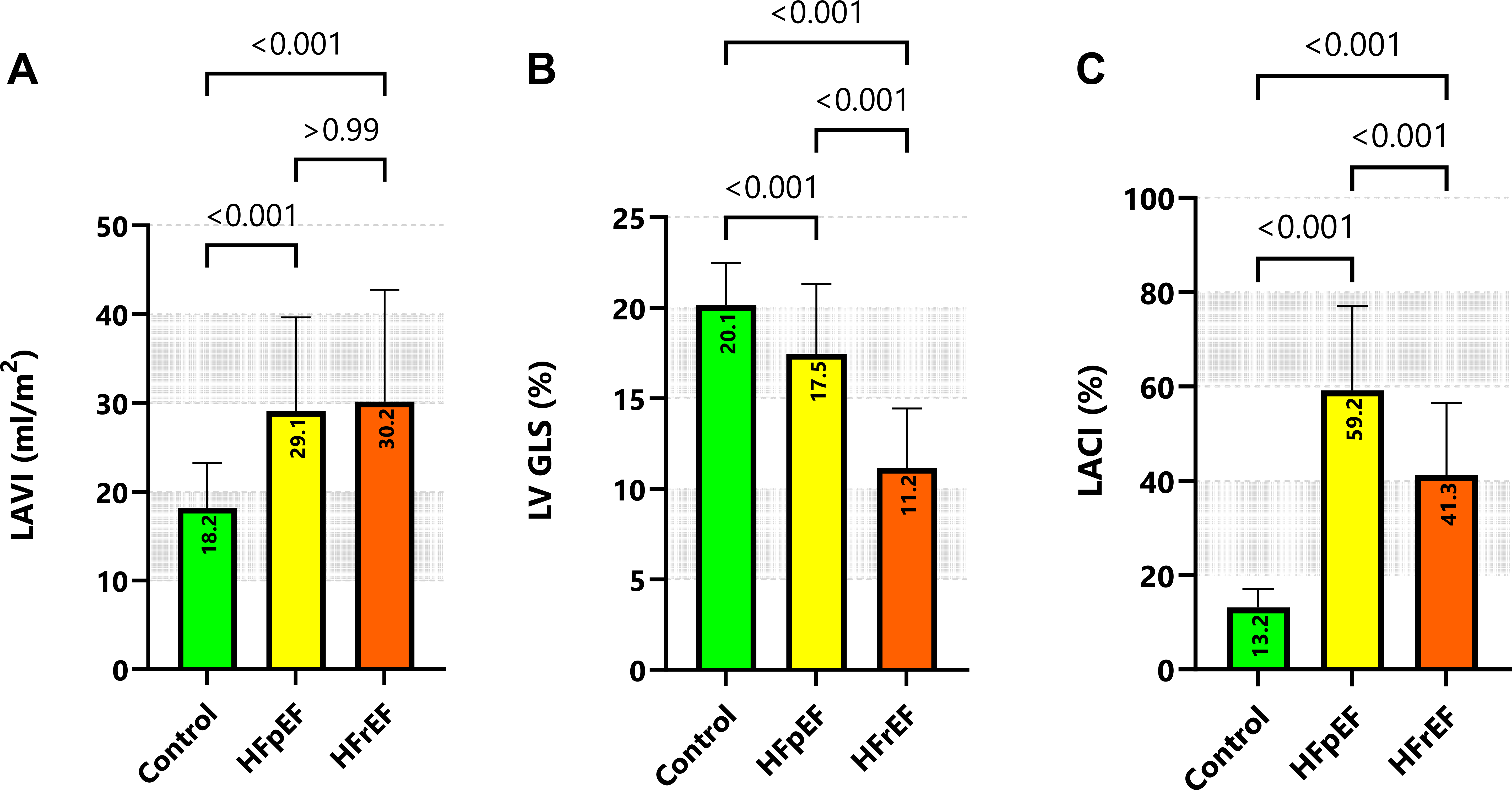
